# Tuberculosis presentation and outcomes in elderly Hispanics from Tamaulipas, Mexico

**DOI:** 10.1101/2023.03.14.23287283

**Authors:** Belinda A. Medrano, Miryoung Lee, Gretchen Gemeinhardt, Javier E. Rodríguez-Herrera, Moncerrato García-Viveros, Blanca I. Restrepo

## Abstract

Old people are at high risk of developing and dying from pulmonary infections like tuberculosis (TB), but they are few studies and particularly in Hispanics. To address these gaps, we sought to identify host factors associated with TB and adverse treatment outcomes in old Hispanics by conducting a secondary analysis of TB surveillance data from Tamaulipas, Mexico (2006-2013; n=8,381). Multivariable logistic regressions were assessed for the elderly (ELD, ≥65 years) when compared to young (YA, 18 to 39 years) and middle-aged adults (MAA, 40 to 64 years). We found that the ELD had features associated with a less complicated TB (e.g. less extra-pulmonary TB, abandoning of treatment or having drug resistant TB), and yet, were more likely to die during TB treatment (adj-OR 3.9, 95% 2.5, 5.25). Among the elderly, excess alcohol use and low BMI increased their odds of death, while diabetes and BCG vaccination were protective. These data suggest that old people share some, but not all the risk factors for adverse TB treatment outcomes, when compared with younger adults. Furthermore, even though old age in itself is an important predictor of death during TB, the elderly are not prioritized by the World Health Organization for latent TB infection screening and treatment during contact investigations. We propose the inclusion of the elderly as a high-risk group in TB management guidelines.

## Introduction

*Mycobacterium tuberculosis* is a bacterial disease that has infected a quarter of the world’s population with non-symptomatic latent infection or progressed to active tuberculosis (TB) disease. In 2020, an estimated 9.9 million people developed TB and 1.5 million died [1]. Latent TB infection and active TB disease are more prevalent in the elderly, i.e., adults over the age of 65 [2-4]. This is relevant as the global elderly population grows from 1 out of 11 in 2019 to 1 in 6 by 2050 [5]. The World Health Organization (WHO) regions of the Eastern Mediterranean, South-East Asia, and Western Pacific already have a significant burden of TB cases in the elderly populations with Taiwan anticipating almost 80% of their new TB cases in the elderly by 2035 [2, 6].

The elderly face additional risks of TB reactivation and infection compared to younger adults, such as compromised immunity, comorbid conditions, and increased TB exposures due to clustering in nursing homes [7-9]. TB and general mortality are highest in the elderly group [2, 10]. An aging population, longer life expectancy, and reactivation of TB disease in the elderly pose challenges to meeting the World Health Organization’s ‘End TB Strategy’ goals of a 90% and 95% reduction in TB incidence and deaths, respectively, by 2035 [1, 8, 10, 11].

Despite their high vulnerability, TB in the elderly is understudied, and the available literature is mostly from Asian populations, with few in Hispanics [12-14]. Studies in Latin American Hispanics are warranted given that risk factors for TB such as poverty, migration, homelessness, incarceration, and diabetes have increased in the last decade [15, 16]. Even in a high-income country like the United States, the TB case rate is nine times higher for Hispanics than for non-Hispanic whites (Centers for Disease Control and Prevention [17].

Given the impact of old age on TB elimination efforts, it is important to study the elderly TB group with the goals of improving patient outcomes and preventing additional TB cases. To address this knowledge gap, and particularly among Hispanics, we aimed to identify sociodemographic and medical risk factors and secular trends for TB in the elderly from the state of Tamaulipas in northeastern Mexico. We also sought to identify the risk factors associated with adverse treatment TB outcomes in old age.

## Materials and Methods

### Study Population

We conducted a secondary analysis of existing TB surveillance data on adults ages 18 to 99 years with a new TB diagnosis and who were reported to the state of Tamaulipas, Mexico between 2006 and 2013. The state’s TB program surveillance dataset was described previously [18]. Briefly, the Tamaulipas’s TB surveillance dataset was comprised of 12,748 TB patients enrolled between 2005 and 2013, and we excluded repeat episodes from the same subject (n=1,827), individuals less than 18 years old (n=624), recurring TB cases (n=777), and individuals enrolled in 2005 who had poor data quality (n=1,089). This left 8,431 new adult TB patients [18]. For this study, we further excluded 43 individuals identified as having a previous TB episode and seven with missing data on TB treatment outcomes, for a final sample size of 8,381 TB patients. Patient data was deidentified and the protocol was approved from the Internal Review Board from UTHealth Houston (HSC-SPH-15-0489) and the Secretaría de Salud de Tamaulipas (076/2015/CEI).

### Participant Characteristics

We compared the elderly group (65 yrs. or older, ELD) to young adults (18-39 yrs., YA) or middle-aged adults (40-64 yrs., MAA). Sociodemographic variables included age, sex, education, Bacille Calmette-Guerin (BCG) vaccination at birth based on the presence of a BCG scar, self-reported excess alcohol use or intravenous (IV) drug use, and low body mass index (BMI <18.5 kg/m^2^ at the time of TB diagnosis). Comorbidities included chronic obstructive pulmonary disease (COPD), self-reported diabetes, and self-reported or laboratory-confirmed HIV. Variables related to TB at the time of diagnosis included disease location (extrapulmonary or pulmonary), positive acid-fast bacilli (AFB) on sputum smears, diagnosis method [culture confirmation by isolation of *M. tuberculosis*, positive AFB smears only, or in their absence, a combination of clinical, epidemiological, radiological and/or histopathological findings], and the number of close contacts. In Mexico, drug susceptibility testing is limited to patients with high suspicion of drug-resistant TB with results available for isoniazid (INH), rifampin (RIF), pyrazinamide (PZA), streptomycin (STR), and ethambutol (EMB). *Mycobacterium tuberculosis* was classified as drug resistant (DR-TB) if resistant to any of the five TB drugs and multi-drug resistant (MDR-TB) when at least INH and RIF were resistant. The adverse outcomes of interest included TB treatment failure (positive AFB smears after five months of TB treatment), abandoned treatment (lost during follow-up for more than three months), and death due to any cause during TB treatment.

### Statistical Analyses

Two-group comparisons of binary variables were analyzed by Pearson’s chi-square or Fisher’s exact test and the Student’s-t test was used for mean comparisons between two groups. Increasing or decreasing trends across age groups or across time were established by the score test for the trend of odds for binary variables. To conduct comparisons between-group, trends across age groups, and trends across time for polytomous and numerical variables, we used the nptrend command in STATA, which is a nonparametric test for trends across ordered groups (an extension of the Wilcoxon rank-sum test). For multiple logistic regressions models, we used backward selection with consideration of variables with p values less than 0.20 in bivariable analysis, and keeping age and sex as key sociodemographics in final models. Effect modification variables for age and each predictor variable were included in simple logistic regression models for adverse outcomes, to determine if age modifies the observed effect of each predictor variable. Statistical significance was set at Type I error (alpha) level <0.05. Data analysis was performed using STATA IC (Stata Corp LLC, College Station, TX) 14.

## Results

### Sociodemographics and Comorbidities in the Elderly

The characteristics of the 8,381 TB patients for analysis are described in **Table 1**. Their mean age was 43 years (SD 16.6; range 18 to 97). Males comprised 65.5% of all TB patients. Less than half (42.2%) had a high school degree or higher education, three-fourths (76.2%) had a history of BCG vaccination, 5.6% reported excess alcohol use, 0.5% were IV drug users, and 8.3% had low BMI. The most prevalent comorbidity was diabetes (25.2%), followed by HIV (5.3%) and COPD (0.9%).

**Table 1.**
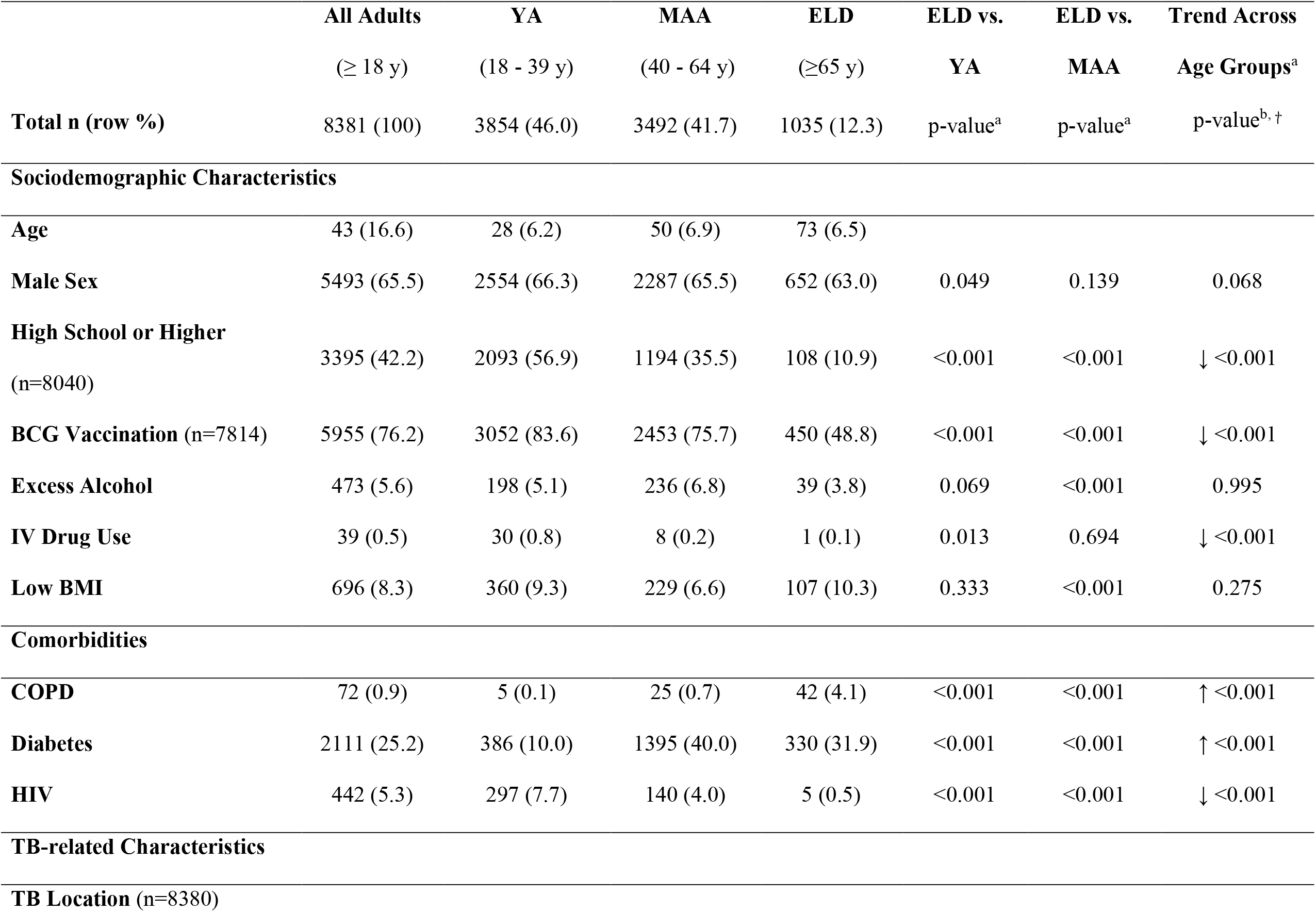
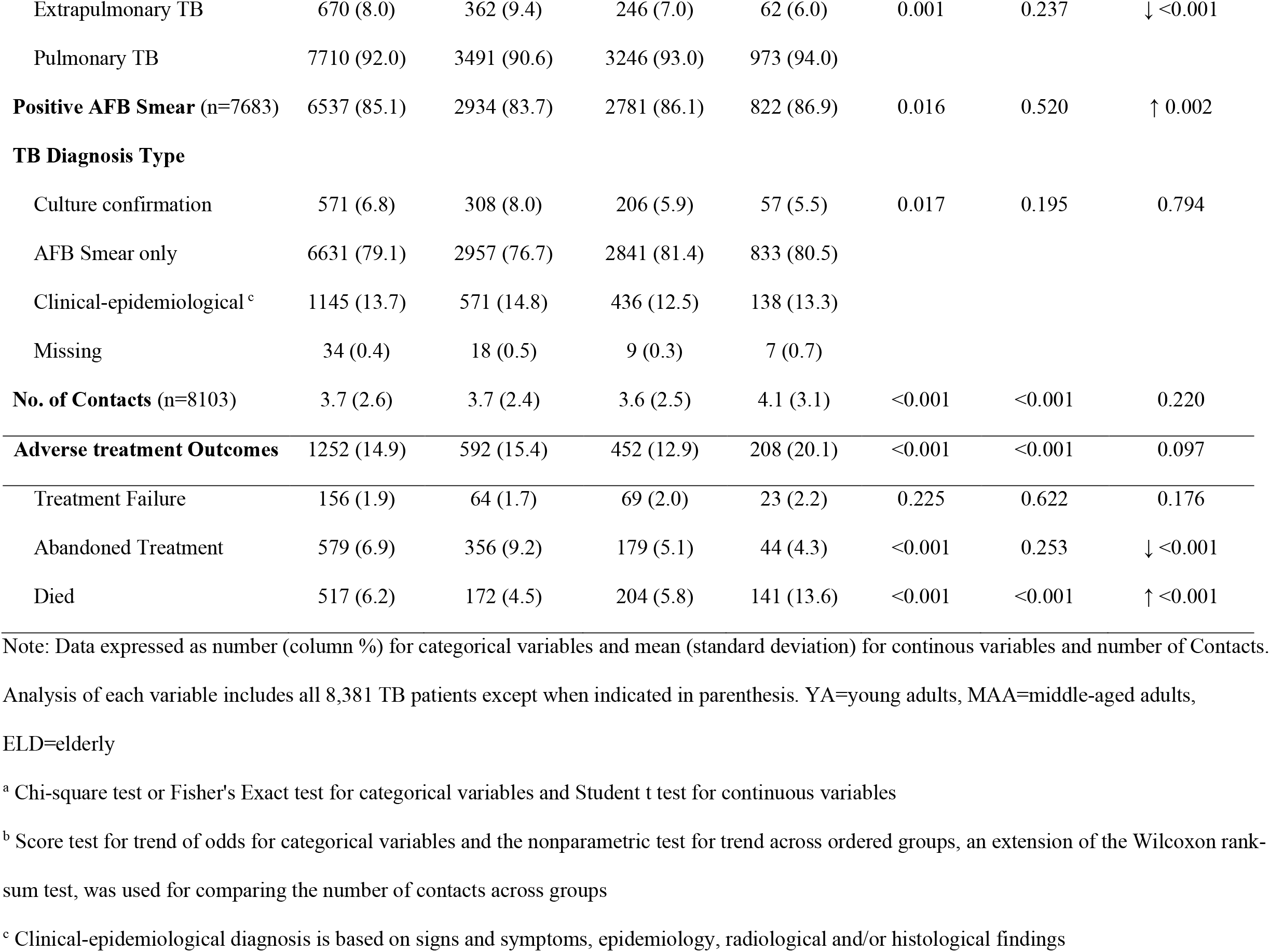
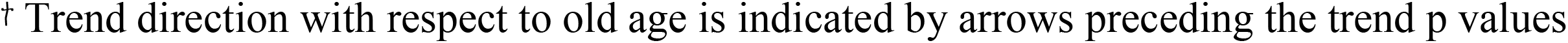
Descriptive Characteristics of TB Patients by Age Group, Tamaulipas 2006 - 2013

The distribution of TB patients by age group was: 46% in YA, 42% in MAA, and 12% in ELD. Mean ages for YA, MAA, and ELD TB patient groups were 28 (SD 6.2), 50 (SD 6.9), and 73 (SD 6.5) years, respectively. The ELD differed from the two younger age groups in their lower level of education (high school degree or higher in 10.9% in ELD vs 56.9% in YA or 35.5% in MAA), BCG vaccination (48.8% in ELD vs 83.6% in YA and 75.7% in MAA), and IV drug use (0.1% in ELD vs 0.8% in YA). Accordingly, significant trends with older age (from YA to MAA to ELD) included a decrease in education level, BCG vaccination rates, and IV drug use, trend p <0.001 for all. Low BMI was highest in the ELD (10.3%) followed by YA (9.3%) and MAA (6.6%).

For comorbidities, COPD was more common among the ELD (4.1% in ELD vs 0.1% in YA and 0.7% in MAA; p <0.001 for increasing trend with older age). Diabetes increased with older age (trend p <0.001), although MAA had the highest prevalence (40%), with the ELD having 31.9% and YA 10% (p <0.001). HIV prevalence was lowest in the ELD (0.5% in ELD vs 7.7% YA and 4.0% in MAA; p <0.001 for decreasing trend with older age).

### TB-related Characteristics

Most patients had pulmonary TB (92%) and a positive AFB smear at the time of diagnosis (85.1%; **Table 1)**. Cultures are not routinely performed in Tamaulipas, and hence, the diagnosis was mostly based on smear results (79.1%), with only 6.8% being culture-confirmed. Each TB patient had an average of 3.7 close contacts. When compared to YA, the ELD had less extra-pulmonary TB (ELD 6.0% vs YA 9.4%; decreasing trend p <0.001) or culture-confirmed TB (ELD 5.5% vs YA 8.0%). The ELD had the highest mean number of close contacts (ELD 4.1 vs 3.7 in YA or 3.6 in MAA).

### TB Drug Resistance

Drug susceptibility testing was only available for 1,165 (13.9%) TB patients, with more testing in YA (15.4%) and MAA (14.2%) vs. the ELD (7.3%; trend p <0.001; **Table 2**). For primary analysis, we assumed that the absence of susceptibility results indicated drug-susceptible TB since testing is prompted when DR-TB is suspected (**Table 2)**. We found that DR-TB decreased with age (trend p=0.030). Among all age groups, the highest monoresistance was for INH (1.2%), and across age groups, only INH monoresistance showed a decreasing trend with age (p=0.002). MDR-TB occurred in 0.6% of all patients and did not differ across age groups. Similar analysis but limited to isolates tested for drug resistance provided similar findings (**S1 Table**).

**Table 2.** TB Drug Resistance Prevalence by Age Group

|  | **All Adults** | **YA** | **MAA** | **ELD** | **Trend across**<br>**age groups** |
| --- | --- | --- | --- | --- | --- |
|  | (≥ 18 y) | (18 - 39 y) | (40 - 64 y) | (≥65 y) | p-value<sup>b,†</sup> |
| **Total n** | 8381 | 3854 | 3492 | 1035 |  |
| **Tested** | 1165 (13.9) | 593 (15.4) | 496 (14.2) | 76 (7.3) | ↓ <0.001 |
| **DR-TB** | 257 (3.1) | 134 (3.5) | 99 (2.8) | 24 (2.3) | ↓ 0.030 |
| **INH** | 99 (1.2) | 57 (1.5) | 39 (1.1) | 3 (0.3) | ↓ 0.002 |
| **RIF** | 6 (0.1) | 4 (0.1) | 2 (0.1) | 0 (0) | 0.238 |
| **PZA** | 6 (0.1) | 2 (0.1) | 3 (0.1) | 1 (0.1) | 0.544 |
| **STR** | 36 (0.4) | 12 (0.3) | 17 (0.5) | 7 (0.7) | 0.083 |
| **EMB** | 4 (0.1) | 3 (0.1) | 0 (0) | 1 (0.1) | 0.633 |
| **MDR-TB** | 54 (0.6) | 26 (0.7) | 22 (0.6) | 6 (0.6) | 0.715 |

*Note:* Data expressed as n (column %). Total n = Total number of TB patients in each age group. This is the denominator for all calculations where those not tested were considered drug-susceptible as described in the text. YA=young adults, MAA=middle-aged adults, ELD=elderly. Tested = susceptibility testing documented. DR-TB= resistant to any of the TB drugs listed. Mono-resistance is listed for individual antibiotics: INH= isoniazid, RIF= rifampin, PZA= pyrazinamide, STR= streptomycin, EMB= ethambutol. MDR-TB=multi-drug resistant TB defined as resistant to at least INH and RIF.
<sup>a</sup> Chi-square or Fisher's Exact test
<sup>b</sup> Score test for trend of odds across the three age groups
<sup>†</sup> Trend direction with respect to old age is indicated by arrows preceding the trend p values

### Adverse TB Treatment Outcomes

Among all patients, 14.9% had one of the following adverse outcomes: treatment failure (1.9%), abandoned treatment (6.9%), or death (6.2%), **Table 1**. Adverse TB treatment outcomes were highest among the ELD (20.1%) followed by YA (15.4%) and MAA (12.9%) groups. Treatment failure did not differ between age groups, but the ELD group was less likely to abandon treatment (4.3%) when compared to YA (9.2%; p < 0.001) with a decreasing trend with older age (trend p <0.001). In contrast, the ELD were more likely to die during TB treatment (13.6%) when compared to YA (4.5%) or MAA (5.8%) with an increasing trend with older age (trend p <0.001).

For multivariable analysis, we examined whether old age was independently associated with adverse TB treatment outcomes with YA as the reference group. IV drug use and HIV were excluded as predictor variables due to their low prevalence in the ELD group (**Table 3**). Age groups were not associated with treatment failure. Instead, among all age groups, adjusted models showed that treatment failure was higher for males, although statistical significance was not reached (adj. OR 1.45, 95% CI 0.99, 2.10), those with positive AFB smears (adj. OR 1.97, 95% CI 1.03, 3.79) or DR-TB, (adj. OR 13.85, 95% CI 9.42, 20.37). Higher education was protective against treatment failure (adj. OR 0.69, 95% CI 0.48, 0.99). Abandoning treatment was 59% lower in the elderly (adj. OR 0.41, 95% CI 0.28, 0.59) when compared to YA (**Table 3**). Among all TB patients, the factors associated with higher odds of abandoning treatment were male sex (adj. OR 2.09, 95% CI 1.65, 2.66), excess alcohol use (adj. OR 1.44, 95% CI 1.03, 2.01), and low BMI (adj. OR 1.59, 95% CI 1.18, 2.13). Protective factors against abandoning treatment included high school or higher education (adj. OR 0.74, 95% CI 0.60, 0.90), having extrapulmonary disease (adj. OR 0.51, 95% CI 0.30, 0.87), and positive AFB smear at diagnosis (adj. OR 0.63, 95% CI 0.49, 0.82). The odds of abandoning treatment decreased by 11% (adj. OR 0.89, 95% CI 0.85, 0.93) for each additional close contact reported by a TB patient. Death was four-fold higher with old age (ELD vs YA: adj. OR 3.90, 95% CI 2.90, 5.25; **Table 3**).

**Table 3.**
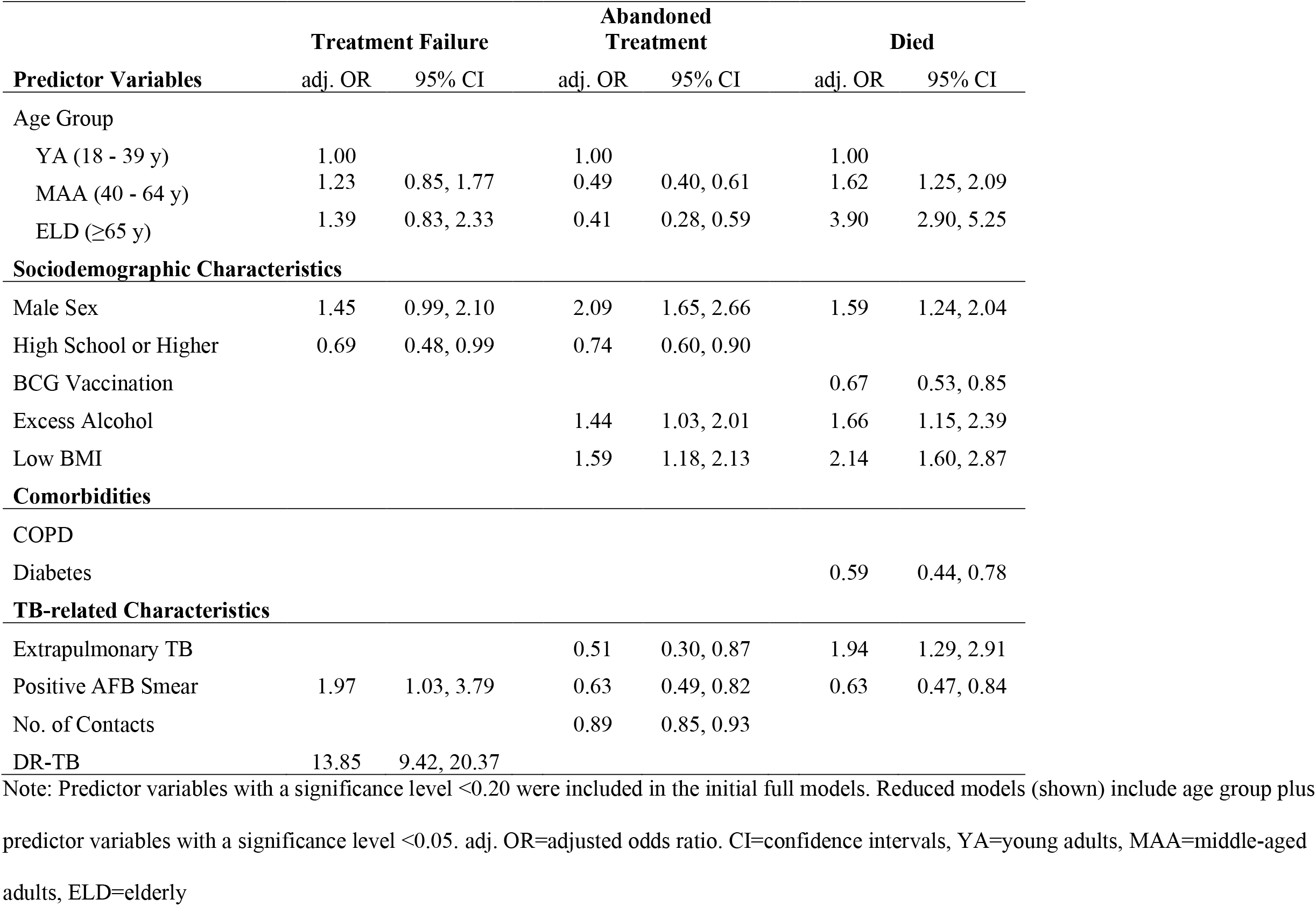
Multivariable analyses to identify whether old age (ELD) is an independent predictor of treatment failure, abandoned treatment, or death

Among all TB patients, the odds of death were also higher for MAA (adj. OR 1.62, 95% CI 1.25, 2.09), males (adj. OR 1.59, 95% CI 1.24, 2.04), excess alcohol users (adj. OR 1.66, 95% CI 1.15, 2.39), those with low BMI (adj. OR 2.14, 95% CI 1.60, 2.87), and extrapulmonary disease (adj. OR 1.94, 95% CI 1.29, 2.91). BCG vaccination (adj. OR 0.67, 95% CI 0.53, 0.85), diabetes (adj. OR 0.59, 95% CI 0.44, 0.78), and positive AFB smear (adj. OR 0.63, 95% CI 0.47, 0.84) were protective against death.

We sought to identify independent predictors of adverse outcomes among the ELD group alone (**S2 Table**). In the ELD, the odds of treatment failure was 14 times higher in patients with DR-TB (adj. OR 14.10, 95% CI 4.69, 42.32). Abandoning treatment decreased by 56% for those with a positive AFB smear and by 15% for each close contact. The odds of death increased by 4% for each additional year of life, and by more than 2-fold in those with excess alcohol use or low BMI. Male sex was not associated with any adverse outcome in the ELD (**S2 Table**), despite its relationship with treatment failure, abandoning treatment and death in all age groups (**Table 3**).

Finally, we evaluated if among all TB patients, age would modify the associations between host factors and adverse TB treatment outcomes. However, multivariable models with additional interaction terms for age did not show significant associations (**S3 Table**).

### Trends Over the Eight-year Period in the ELD

We evaluated if there were trends over time (2006 --2013) in the prevalence or characteristics of the ELD group. Among all TB patients, the proportion of ELD remained similar across the years (range 11.2 – 13.8%; p 0.531; **S4 Table**). Among the ELD group only, there was an increasing trend in education level (p <0.001), BCG vaccination (p <0.001), diabetes (p 0.010), extrapulmonary TB (p 0.005) and DR-TB (p 0.002; **Tables 4 and S4**). We also evaluated trends over time for younger adults (**Table 4**) and found that unlike the ELD, the YA and MAA had a reduction in excess alcohol use, low BMI, and abandoning of TB treatment, and that the YA had a decreasing trend in death.

**Table 4.**
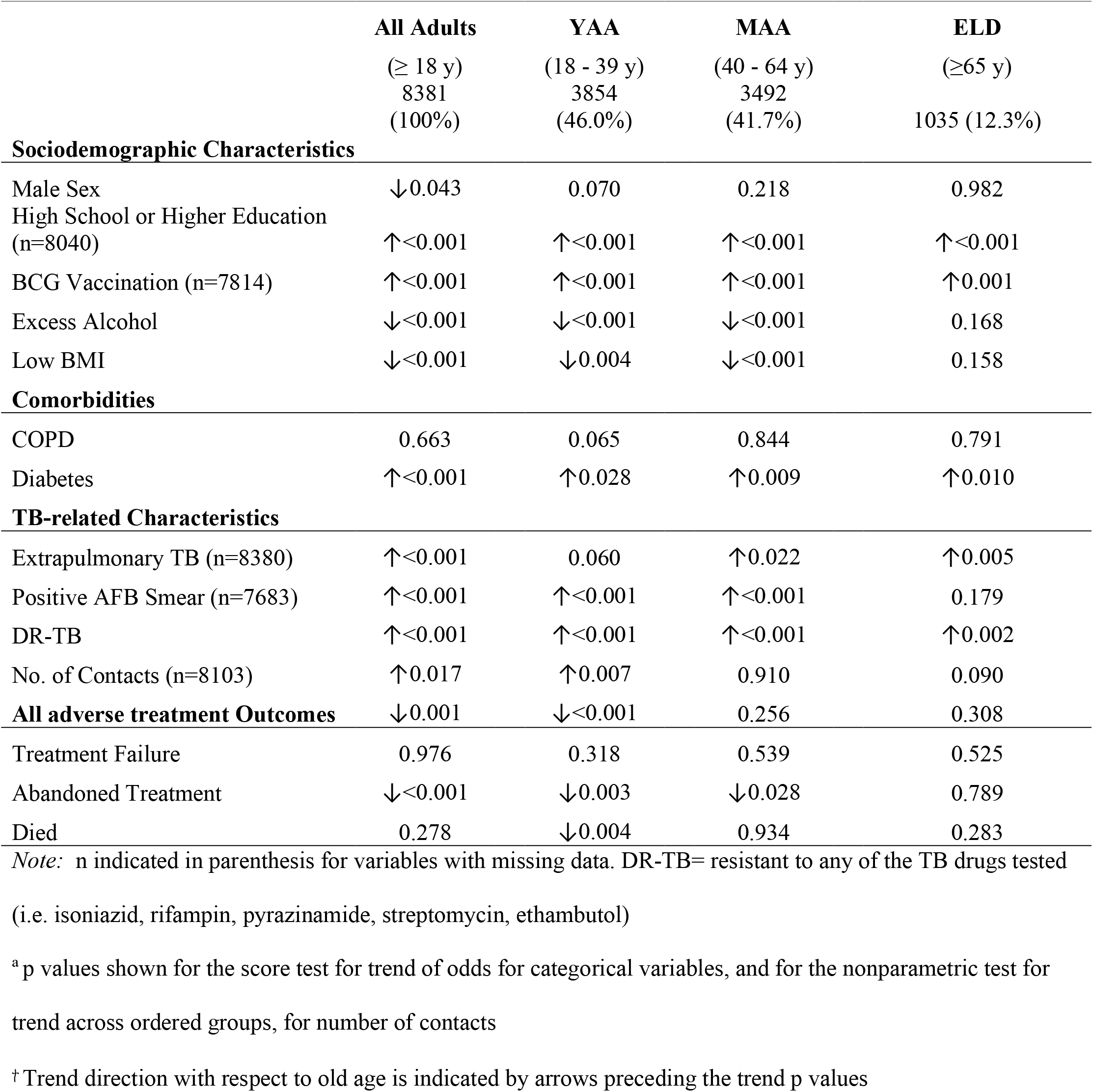
Trends across time (2006--2013) in sociodemographic and clinical characteristics of TB patients by age groups a, ^†^

|  | **All Adults**<br>(≥ 18 y)<br>8381<br>(100%) | **YAA**<br>(18 - 39 y)<br>3854<br>(46.0%) | **MAA**<br>(40 - 64 y)<br>3492<br>(41.7%) | **ELD**<br>(≥65 y)<br>1035 (12.3%) |
| --- | --- | --- | --- | --- |
| **Sociodemographic Characteristics** |  |  |  |  |
| Male Sex | ↓0.043 | 0.070 | 0.218 | 0.982 |
| High School or Higher Education<br>(n=8040) | ↑<0.001 | ↑<0.001 | ↑<0.001 | ↑<0.001 |
| BCG Vaccination (n=7814) | ↑<0.001 | ↑<0.001 | ↑<0.001 | ↑0.001 |
| Excess Alcohol | ↓<0.001 | ↓<0.001 | ↓<0.001 | 0.168 |
| Low BMI | ↓<0.001 | ↓0.004 | ↓<0.001 | 0.158 |
| **Comorbidities** |  |  |  |  |
| COPD | 0.663 | 0.065 | 0.844 | 0.791 |
| Diabetes | ↑<0.001 | ↑0.028 | ↑0.009 | ↑0.010 |
| **TB-related Characteristics** |  |  |  |  |
| Extrapulmonary TB (n=8380) | ↑<0.001 | 0.060 | ↑0.022 | ↑0.005 |
| Positive AFB Smear (n=7683) | ↑<0.001 | ↑<0.001 | ↑<0.001 | 0.179 |
| DR-TB | ↑<0.001 | ↑<0.001 | ↑<0.001 | ↑0.002 |
| No. of Contacts (n=8103) | ↑0.017 | ↑0.007 | 0.910 | 0.090 |
| **All adverse treatment Outcomes** | ↓0.001 | ↓<0.001 | 0.256 | 0.308 |
| Treatment Failure | 0.976 | 0.318 | 0.539 | 0.525 |
| Abandoned Treatment | ↓<0.001 | ↓0.003 | ↓0.028 | 0.789 |
| Died | 0.278 | ↓0.004 | 0.934 | 0.283 |

*Note:* n indicated in parenthesis for variables with missing data. DR-TB= resistant to any of the TB drugs tested (i.e. isoniazid, rifampin, pyrazinamide, streptomycin, ethambutol)
<sup>a</sup> p values shown for the score test for trend of odds for categorical variables, and for the nonparametric test for trend across ordered groups, for number of contacts
<sup>†</sup> Trend direction with respect to old age is indicated by arrows preceding the trend p values

## Discussion

Despite projections showing a worldwide increase in the proportion of older individuals, studies in the elderly are underrepresented when compared to those in younger adults [19-21]. This knowledge gap extends to the elderly with TB, especially in the Americas. To address this disparity, we sought to obtain a better understanding of elderly Hispanics with TB using TB surveillance data from the Mexican state of Tamaulipas. The elderly comprised 12.3% of all adult TB patients, and over the eight-year study period we observed: i) an improvement in factors that could be protective for TB (e.g. higher education and BCG vaccination coverage), ii) an increase in factors that favor adverse TB outcomes (e.g. extrapulmonary TB and DR-TB), and iii) an increase in diabetes, which had a protective effect on TB outcomes in our ELD cohort as discussed below [22-26]. The elderly differed from younger adults in their higher prevalence of COPD, which could increase their odds of a more severe TB disease, and in having less BCG vaccination and lower education level, which have been associated with higher odds of TB [14, 27, 28]. However, old age was also associated with factors known to improve their TB prognoses such as less IV drug use, HIV, and extra-pulmonary TB, and abandonment of treatment [29-31]. The distribution of risk factors for TB, therefore, can have variations across all adult age groups and identifying the high-risk profiles in the elderly is important for prompt TB suspicion, diagnosis, and treatment initiation, in order to improve their prognosis.

Regardless of a combination of risk and protective factors for TB in the elderly in multivariable models, the highest threat to this age group was their higher odds of death. This finding is consistent with previous studies in the elderly, and we now report it in our Hispanic TB patients [9, 32]. And among the elderly with TB, death was associated with an increase in age, excess alcohol use and low BMI. These factors also contribute to the higher odds of death in all adult age groups [1, 2, 25, 33, 34]. In contrast, male sex was a risk factor for death in YA and MAA TB patients, but not in the ELD in our study.

The prevalence of type 2 diabetes increases with age, as observed in our TB cohort [35, 36]. Diabetes is a risk for TB in adults, this includes findings using prospective or retrospective cohorts from Hispanics in US and Mexican communities [18, 37, 38] but not in older people [13, 14]. We posit that differences between diabetes in younger versus older adults may be explained by differences in the pathophysiology and/or the management of type 2 diabetes [13]. While diabetes in the elderly may not be a risk factor for TB development, in the present study, we evaluated if it would be associated with higher odds of death. We found the opposite, with diabetes being protective, as reported in Brazil by [39]. A higher risk of adverse TB treatment outcomes in the presence of diabetes has been reported in previous studies, although not specifically evaluated in older patients [9, 25, 40]. A possible explanation for these discrepant findings may be the higher prevalence of obesity in diabetes patients. While diabetes is a risk factor for TB, obesity is protective against TB [41, 42]. It is possible that the higher prevalence of obesity in our study population offsets, to some extent, the risk of death during TB that is conferred by diabetes [43]. Consistent with this finding, a study in adults from India reported that diabetes offsets the risk of adverse outcomes conferred by low BMI [44]. Further studies are needed to understand the relationship between diabetes, BMI, death in TB patients, and old age.

We found that BCG vaccination at birth decreased with older age. This is consistent with the lower prevalence of BCG vaccination in the elderly versus younger adults with TB in a prospective research cohort [14]. In that study, BCG vaccination at birth was associated with lower odds of TB, but only in the elderly. This finding was intriguing given that BCG is known to protect infants and young children from TB, but not adults [1]. Here we find that BCG vaccination at birth is associated with lower odds of death from TB in all age groups. Our findings are consistent with the association between unfavorable TB treatment outcomes and absence of a BCG scar in a Malaysian population [25]. Together, these findings suggest a possible protective role of BCG with respect to TB risk or death, although further studies are needed to rule out confounding factors that were not available in the surveillance dataset, such as lower access to healthcare in non-BCG-vaccinated individuals. At the cellular level, BCG vaccination induces epigenetic changes in innate immune cells (e.g., trained immunity) that associate with protection against TB and other microbial infections [45]. Thus, basic science studies are also warranted to help elucidate how BCG vaccination at birth may contribute to TB protection and lower the risk of death in the elderly.

We found a decreasing prevalence of extrapulmonary TB with older age, which is consistent with previous studies [31, 46]. These age-related differences may be due to a lower prevalence of comorbidities like HIV, when compared to younger TB patients, that favor extrapulmonary disease [31, 46, 47].

Abandonment of treatment contributes to adverse outcomes but we found that the ELD are less likely to abandon TB treatment [30, 48, 49]. Interestingly, we found an inverse association between the number of TB contacts and odds of abandoning treatment in all age groups, or among ELD patients only. Further studies are needed to determine if this finding is consistent with reports indicating that the level and quality of social support received by patients affects their health outcomes and adherence to treatments [50-53].

Patients with drug-resistant TB are more likely to have adverse treatment outcomes [1, 49, 54, 55]. While we found a lower prevalence of DR-TB in the ELD, our findings point to the importance of identifying these patients given their higher odds of treatment failure.

Our large sample size, coverage over an eight-year period, and breakdown of age into three groups, allowed for an in-depth cross-sectional assessment of unique features in ELD TB patients with respect to their sociodemographics, comorbidities, TB characteristics, and treatment outcomes. Limitations included the secondary nature of the data analyses. Diabetes diagnosis and duration of disease, excess alcohol use, and, to some extent, HIV status were self-reported and may be underestimated, although we do not anticipate that there would be a differential underreporting of these host characteristics in the ELD when compared to other age groups. Low BMI was reported as a categorical variable with no further granularity to assess the contribution of different levels of body weight or obesity to TB risk or protection in the elderly. Smoking is a risk factor for TB and known to be more prevalent in older cohorts, but not reported in this surveillance dataset [14]. Finally, DR-TB testing was only evaluated upon suspicion, as in most TB-endemic countries worldwide, and hence, pan-susceptibility was assumed when data was not available. However, analysis limited to patients in whom drug susceptibility was tested provided similar results (**Table S1**).

Our study raises awareness of the high vulnerability of older people to death from TB. Per WHO’s latent TB infection management guidelines, TB contact investigations focus on the identification of high-risk groups for TB development. However, these are limited to children below 5 years of age or people living with HIV, and of other adults and children household contacts in low TB incidence countries without mention of the growing elderly population [56]. Our findings call for the need to consider the elderly in a similar high-risk category. A recent study showed that Interferon-gamma release assays (IGRA) are effective for identification of latent TB infection in the elderly [57]. We also identified modifiable sociodemographic factors associated with higher odds of death during TB in the elderly, such as excess alcohol use or low BMI. Close supervision of elderly with new TB or LTBI should be offered for counseling of alcohol use or nutritional support. We also found that abandoning treatment was more likely when patients have fewer contacts. Hence, protocols to promote social workers at TB clinics to reach out to family members to emphasize the importance of social support, can have a positive impact on treatment success. By studying the elderly TB group separately from the other distinct adult age groups, we will have a better understanding of how to approach the challenges of TB eradication in this understudied and vulnerable population.

## Data Availability

Original datasets used for analysis of the findings presented in this manuscript are available upon request.

## Acknowledgements

We thank the personnel from all the sanitary jurisdictions of the Secretaría de Salud de Tamaulipas who contributed to the collection and recording of data from the TB patients, and Dr. Santa Elizabeth Ceballos Liceaga for providing the surveillance datasets for our analysis as coordinator of the Sistema de Vigilancia Epidemiológica de Tuberculosis y Lepra, Dirección General de Epidemiología, Secretaría de Salud de Mexico. We also thank Dr. Bassent Abdelbary for generation of the basic dataset used for analysis and guidance for data analysis.

## Supporting Information

**S1 Table. TB drugs susceptibility sub-analysis that excludes not tested and unknown from drug resistance calculations**.

**S2 Table. Multivariable analyses for predicting treatment failure, abandoned treatment, and death in the elderly (ELD) group**.

**S3 Table. Evaluation of age as an effect modifier of the association between the predictor variables listed and adverse TB outcomes**.

**S4 Table. Trends over time (2006 – 2013) in the prevalence of the Elderly group or their characteristics**.

